# Assessment of Post COVID-19 Health Problems and its Determinants in North India: A descriptive cross section study

**DOI:** 10.1101/2021.10.03.21264490

**Authors:** Suraj Singh Senjam, Yatan Pal Singh Balhara, Parmeshwar Kumar, Neeraj Nichal, Souvik Manna, Karan Madan, Nishat Hussain Ahmed, Noopur Gupta, Rajesh Sharma, Yashdeep Gupta, Animesh Ray, Vivek Gupta, Praveen Vashist, Atul Kumar, Lalit Dar, Jeevan Singh Titiyal, Radhika Tandon, Randeep Gulleira

## Abstract

With millions of people getting affected with COVID-19 pandemic caused by a novel severe acute respiratory syndrome corona virus 2 (SARS-CoV-2), people living with post COVID-19 Symptoms (PCS) are expected to rise in the future· The present study aimed at assessing PCS comprehensively and its associated factors among COVID-19 recovered adult population in north India.

**Methods:** In a tertiary health centre at Delhi, an online based cross-sectional study was conducted using a semi-structured questionnaire, developed by employing a nominal group technique, in aged 18 years and above who were SARS-CoV-2 positive during the month of January to April 2021. Socio-demographic, various potential risk factors, including pre-existing morbidities, vaccination status, and severity of acute COVID-19 illness, information on acute illness for management and a spectrum of PCS were collected between June 16 to July 28, 2021. Each participant was contacted telephonically before sending the survey link. PCS were presented as relative frequency; chi-square test, odds ratio, including adjusted, were calculated to rule out association between PCS and potential predictors.

**Results:** A total of 773 of 1801 COVID recovered participants responded to the link reaching a participation rate of 42·9%, with a median age of 34 years (IQR 27 to 44). Male respondents were 56·4%. Around 33·2% of them had PCS at four or more weeks, affecting almost all body organ systems. The most prevalent PCS were fatigue (79·3%), pain in the joins (33·4%), muscle (29·9%), hair loss (28·0%), headache (27·2%), breathlessness (25·3%), sleep disturbance (25·3%) and cough (24·9%). The prevalence of PCS was reduced to 12·8% at 12 weeks after positive test. Factor such as female gender, older age, oxygen supplementation during the acute illness, working in healthcare care facilities, the severity of acute illness, and pre-existing co-morbid were risk factors for PCS. Further, vaccination (second dose) reduced the odds of developing PCS by 45% compared to unvaccinated participants (aOR 0·65; 95%CI 0·45-0·96). Finally, 8·3% of participants rated their overall health status was either poor or very poor following COVID-19 illness.

**Conclusions:** The PCS involves almost all organ systems, regardless of the severity of acute COVID-19 illness. Two doses of vaccine help to reduce development of PCS.

**Research in Context:** *Evidence before this study:* Although the evidence is mounting in prolonged COVID-19 symptoms among COVID-19 survivors, to date, the full range of such post-COVID-19 symptoms (PCS) is not yet fully understood. There is a lack of studies that assessed PCS comprehensively among persons who have recovered from the COVID-19illness. For example, limited data are available on psychosocial, behavioral, and oral manifestations related to PCS. Further, there is a paucity of studies that included a wide range of determinants of PCS and the association of vaccination with the development of PCS across the world. Our study is the first such study conducted among COVID-19 recovered persons who with a majority of them employed in a tertiary health care institute of north India.

*Added value of this study:* Our study, for the first time, investigated a wide range of post-COVID-19 manifestations among COVID-19 recovered persons in organ-specific and psychosocial behavioral aspects, making this the largest categorization of PCS currently (in total 16). The study included telephonic calls to each eligible candidate which helped in ensuring the COVID-19 status at the time of the study. Since the participants either were employees in the hospital or their dependents that enhance the accuracy of reporting PCS. The most prevalent symptom was unspecific PCS (85.6%), e.g., fatigue, followed by musculoskeletal manifestations (49·8%), Ear, Nose and Throat symptoms (47·5%), neurological (47·0%), cardio-respiratory (42·4%, gastrointestinal (36·2%), ocular symptoms (31·9%), dermatological symptoms (31·5%), and cardio-vascular (24·5%) symptoms, and mental health symptoms (23·7%). The rest of the organ specific symptoms were observed in less than 20% of the respondents. Older age, female gender, pre-existing co-morbid, oxygen supplementation during acute illness, the severity of illness, working in health care institutions were associated with PCS. Vaccination after the second dose was protective against PCS compared to non-vaccinated participants. Further, our study also reported a rating of the overall health status among COVID survivors, whereby around 8.3% of them reported being a poor or very poor health.

*Implications of all the available evidence:* PCS affects a multi-organ organ system, irrespective of the severity of acute-phase COVID-19 illness and hospitalization. Such persistent COVID-19 symptoms, compounded by its heterogeneity among COVID survivors can pose a substantial burden to the affected individuals and their families and additional challenges for healthcare delivery and public health service. The current study shows that one in three individuals experience persistent COVID-19 symptoms. Since the COVID pandemic is still ongoing across the world, therefore, the number of people experiencing PCS is likely to be increased substantially further. An integrated PCS care strategy, but not limited to organ-specific healthcare disciplines, others such as psychosocial support, including counseling and education, rehabilitation, community-based rehabilitation programs will be required for management. Prioritization of PCS care to elder and co-morbid patients should be recommended. Expediting the vaccination drive will be helpful to reduce the development of persistent COVID-19 symptoms. Research, collaborative and multidisciplinary, is required to understand the underlying pathophysiology mechanism for PCS.

## Introduction

The World Health Organization declared the COVID-19 a pandemic on March 22, 2020, around three months after the first case of disease due to the novel coronavirus.^1,2^ Since then, the disease continues to spread in an unprecedented manner across the world causing loss of millions of lives. As of September 18, 2021, more than 226 million people were affected and nearly 5 million people lost their lives due to COVID-19.^3^ In India alone, around 35 million have been infected with the virus and more than 440 thousands people have died due to the disease.^4^

While the countries continue to grapple with the rising number of COVID-19 cases, there is growing evidence on lingering COVID-19 symptoms and health problems extending up to a few months from the initial diagnosis in those who were declared as recovered either clinically or microbiologically. Multiple recent studies, including systemic reviews, illustrated that around 50 to 87% of hospitalized patients experienced at least one or more post COVID-19 symptoms for several weeks even after convalescence or discharge from the hospital.^5678-9^ Around 20% of patients diagnosed with COVID-19 and having symptomatic acute phase reported persistence of symptoms lasting more than 3 months, albeit at a reduced intensity.^10^ Such persisting and debilitating symptoms following COVID-19 mean that the adverse consequences of the pandemic do not end with the recovery from the acute phase of the disease and the pandemic is likely to have ongoing impact on the individuals, family, social, and country as a whole.^5^

As the pandemic continues to spread globally, more people will continue to getting infected. Consequently, the number of people living with post COVID-19 sequelae and symptoms will increase over time. Although the natural history of COVID-19 is not completely known, now it is well recognized that COVID-19 is a multi-organ system disease with a broad spectrum of manifestations.^5^ With millions of individuals recovering, the consequence of post COVID-19 symptoms are likely to become an additional burden on the health care delivery system. Moreover, the long term sequalae of COVID-19 infection are not yet fully known, and hence hamper the attempts to prepare the health care delivery systems to manage the same effectively in the coming months.

Understanding the post COVID-19 symptoms and associated risk factors is crucial to reorient the health infrastructure to make it more responsive to the needs of a large number of persons who are likely to require such medical interventions going ahead. Such information can be used to guide the development of appropriate infrastructure and manpower thereby designing comprehensive post COVID-19 management strategies and patients care plan in hospital as well as in the rehabilitation facilities. Additionally, an appropriate planning will also help with long-term monitoring and follow up. Therefore, we planned the present study that aimed at assessing post COVID-19 health problems and its determinants among patients aged 18 years and older who had recovered from the disease.

### Materials and methods Study population

The present cross-sectional study was conducted at a tertiary health care institute in Delhi, India. The participants were beneficiaries under the Employee Health Insurance Scheme (EHS) of the Institution who were aged 18 years and above. The EHS beneficiaries include not only currently working or retired staff of the institution, but also their respective dependents. All Institutional EHS beneficiaries are entitled for free healthcare services, including COVID-19 related investigation and management.

A list of individuals who have tested positive for SARS-CoV-2 between January 1, 2021 to April 30, 2021 was secured from the Department of Hospital Administration of the institution. We did not procure the data beyond this timeframe. The information available included age, gender, date of diagnostic test for novel coronavirus infection, and their contact details. This list included a total of 2464 patients. We excluded 427 individuals from the study as they were below 18 years of age. The final list included 2037 participants who were approached for participation in the current study.

### Study definitions

Post COVID-19 symptoms (PCS): Symptom(s) that persisted beyond four weeks from the date of SARS-CoV-2 positive test conducted using either Reverse Transcriptase Polymerase Chain Reaction (RT-PCR) or Cartridge Based Nucleic Acid Amplification Test (CB-NAAT). Further, post COVID-19 symptoms were classified as acute or long according to timeframe.

1. Short term post COVID-19 symptoms (ST-PCS): Symptoms present beyond four weeks after the SARS-CoV-2 positive test and lasting less than or up to twelve weeks.
2. Long term post COVID-19 symptoms (LT-PCS). Symptoms present beyond twelve weeks after SARS-CoV-2 positive test.

### Study tool

A semi-structured questionnaire was developed for the study purpose. The questionnaire was digitized using Google forms.^11^ Google forms is one of the online data collection tools that has commonly being used for electronic data collection and surveys in other studies.^12,13^

A nominal group technique was used to develop the questionnaire. The researcher developed first draft of the study tool after extensive review of the relevant published papers to ensure that the tool was comprehensive, and it included all possible post COVID-19 symptoms. One expert each was requested from the departments of Endocrinology, Medicine, Psychiatry, Ophthalmology and Pulmonary Medicine, and of the institution. The questionnaire was revised based on the feedback received from the experts during the group discussions (in person and online). The questionnaire included demographic information (age, gender, blood groups, weight (in kilograms), height (in centimetres), occupation), possible determinants of post COID-19 symptoms (smoking, alcohol consumption, severity of COVID-19, pre-existing health problems, vaccination status before SARS-CoV-2 infection, place of management, oxygen supplementation, if treated in hospital). The final section of the questionnaire consisted of a list of possible COVID-19 symptoms which were categorized according to the various organ systems. Pre-testing of the questionnaire was done among ten persons who had recovered from the COVID-19 for standardization purpose. The data from these persons was not included in the final analysis. After the finalization of the questionnaire in English language, it was translated into the Hindi by a translator. The questionnaire was back translated into the English to ensure the accuracy of the translation by a different expert.

### Data collection

The data was collected between June 16, 2021 and July 30, 2021. We intended to keep a gap of a minimum of 4 weeks between the date of data collection and time since tested conducted for any participant. Before sending the survey link, all participants were contacted telephonically and enquired about the status of their COVID-19 disease. Neither we enquired any SARS-CoV-2 test results during the calls nor in the tool. This was done with an aim to exclude the EHS beneficiaries that had not recovered from acute COVID-19 illness. The potential participants were explained about the study during the telephonic call. Those who expressed interest in participation in the study were sent the link to the survey using WhatsApp platform. If any participant did not install or has the WhatsApp to the number we called, then we requested to provide an alternative mobile number that has WhatsApp in use. This was done to enhance the participant rate.

WhatsApp is a popular medium for communication and has been used in many studies as an electronic platform for communication in various studies. ^14,15,16^ Once we forwarded the survey link, we requested to the participants to save the sender number first, to activate the survey link. The link to the survey opened the Participant Information Sheet (PIS), both in Hindi and English languages. Prior to inclusion in the survey the participants were required to offer informed consent. Only those who offered consent were able to access the survey questions. The participants were free to withdraw from the study at any time without specifying any reason. A total of three reminders at a gap of one to two days were sent to participants who did respond to the survey link sent to them earlier.

### Ethics clearance

The study was approved by the institute ethics committee at the institution. The participants were asked for the digital informed consent at the time of registration. The study was done in accordance with ethical principles of the Declaration of Helsinki.

### Data management and analysis

The responses were automatically collected in a Google spreadsheet linked to the data collection form. The data were analysed using the STATA version 15 (StataCorp 2015, Stata Statistical Software: Release 15, College Station, TX: StataCorp LP). Since, the study population were confirmed cases for COVID-19 disease in the past, the participants were divided into two groups as participants with post COVID-19 symptoms and participants without symptoms for further analysis.

Descriptive analysis was done to summarize the characteristics of the study participants. The data was tested for normality distribution. Continuous variables were presented as mean and standard deviation for normal distribution whereas median and interquartile range were used for variables that were not distributed normally. Post COVID-19 manifestations were presented as relative frequency. Further, the post COVID -19 symptoms were categorized into 15 different groups according to different organ specific manifestations, including a separate category as unspecific post COVID-19 symptoms (UPCS).

Besides, the post COVID-19 symptoms (PCS) were classified into the short term (ST-PCS) and long-term post COVID-19 symptoms (LT-PCS). The duration of symptoms present was estimated from the date of confirmed test and date of responses by the participants. Categorical data were presented as absolute counts, percentage and further compared using chi square, fisher’s exact test. The continuous data were compared using t-test. Univariable and multivariable logistic regression model were used to explore associated explanatory variables with post COVID-19 symptoms. The results were presented as odds ratio and adjusted odds ratio (OR and aOR) along with 95% Confidence Interval (CI). Variables which were significant at p <0·10 in the univariable regression analysis were included in multivariate model. Since it was web-based survey, we followed the CHERRIES checklist guidelines while reporting the data.^17^

## Results

### Characteristics of the study population

Of the 2037 individuals who tested positive for SARS CoV-2 during the months of January to April 2021, 236 participants either refused to participate or could not be contacted due to various reasons such as no responses to our calls, wrong or missing phone numbers, and not being reachable over the mobile phone. Therefore, the survey link could not be shared with them. Finally, we were able to send the survey link to 1801 eligible participants.

Twelve of the 1801 eligible participants did not offer consent to participate in the study. A total of 773 individuals offered consent and filled out the survey questionnaire, reaching the completion rate of 42·9% (Table 1).

**Table 1:**
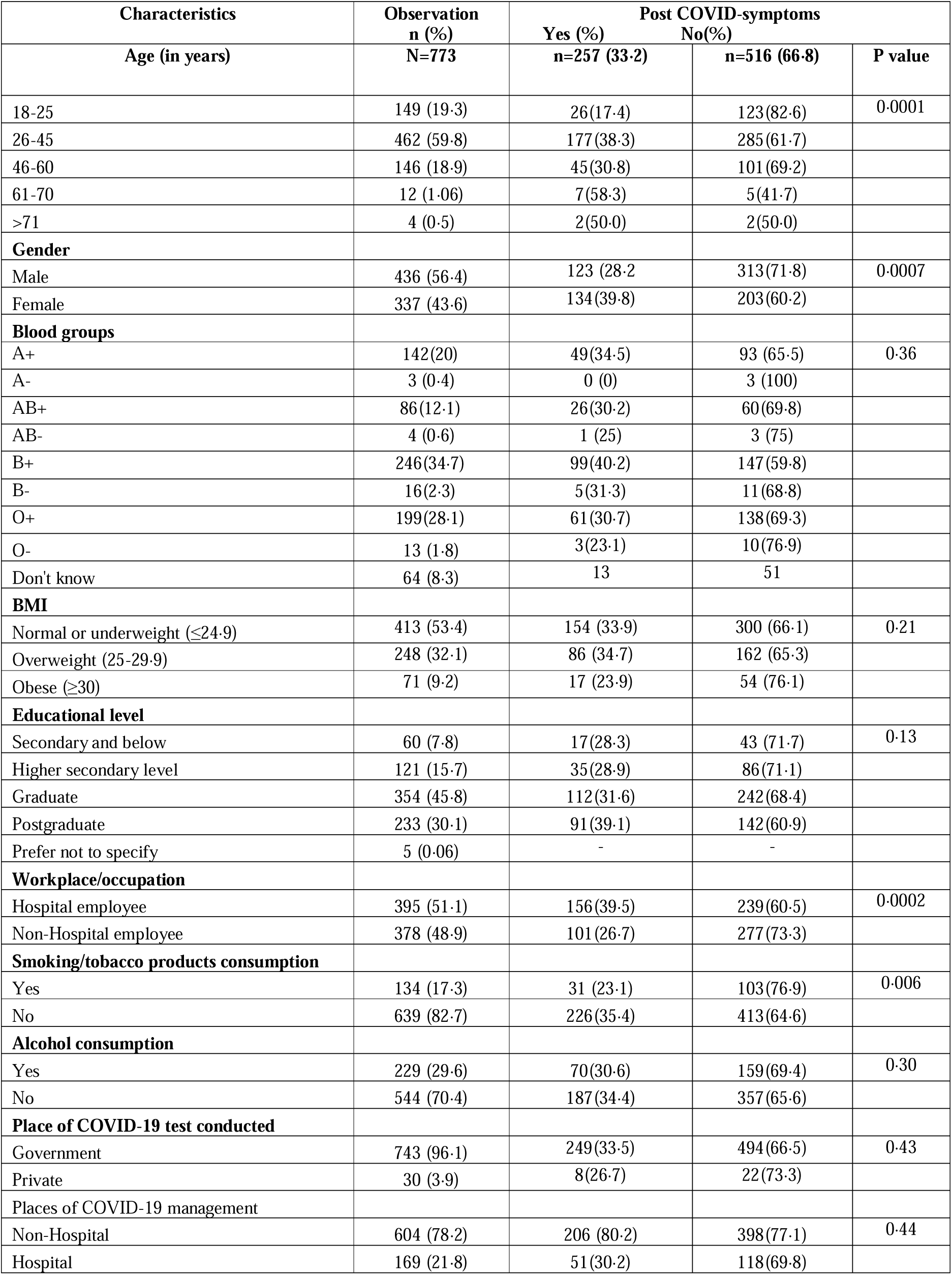
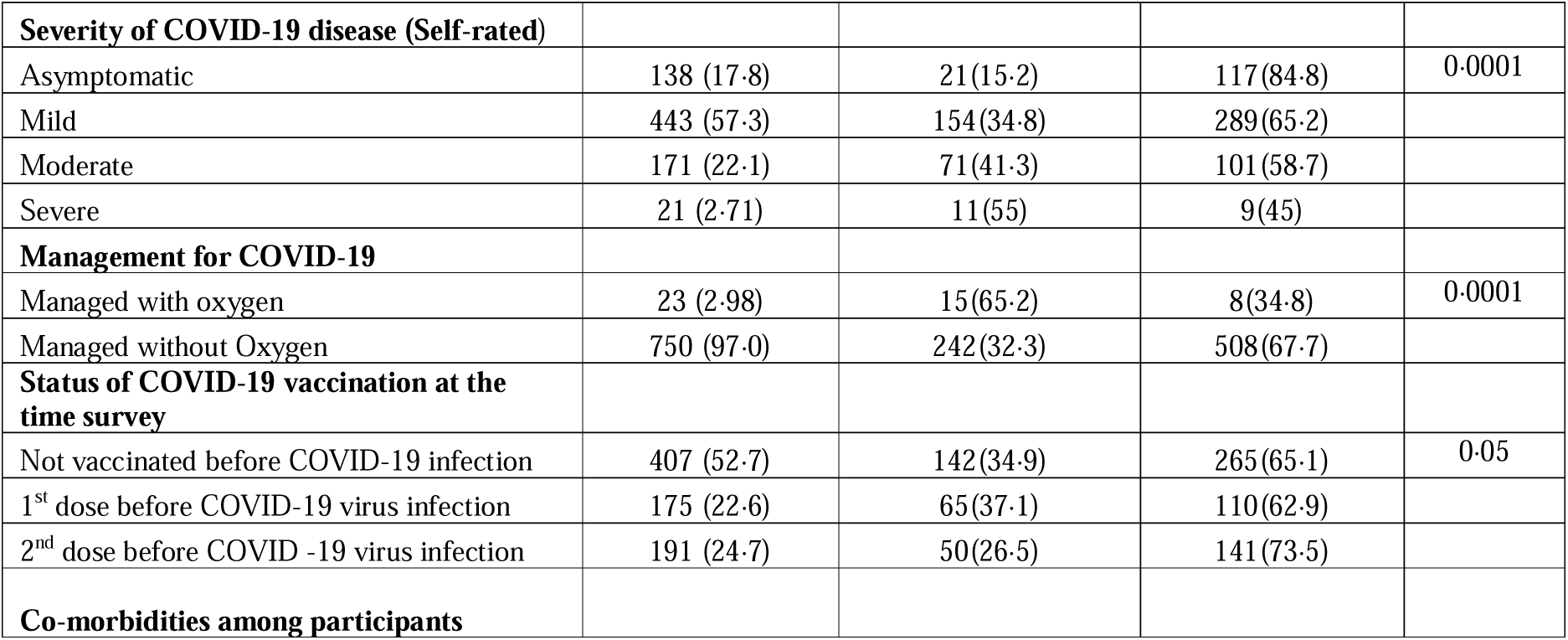
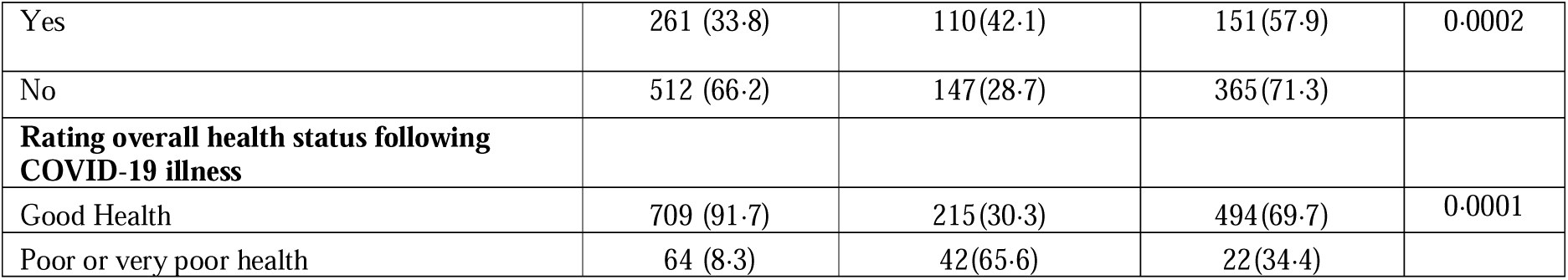
Characteristics of study population.

| Characteristics | Observation<br>n (%) | Post COVID-symptoms |  |  |
| --- | --- | --- | --- | --- |
|  |  | Yes (%)<br>n=257 (33·2) | No(%)<br>n=516 (66·8) | P value |
| **Age (in years)** | **N=773** |  |  |  |
| 18-25 | 149 (19·3) | 26(17·4) | 123(82·6) | 0·0001 |
| 26-45 | 462 (59·8) | 177(38·3) | 285(61·7) |  |
| 46-60 | 146 (18·9) | 45(30·8) | 101(69·2) |  |
| 61-70 | 12 (1·06) | 7(58·3) | 5(41·7) |  |
| >71 | 4 (0·5) | 2(50·0) | 2(50·0) |  |
| **Gender** |  |  |  |  |
| Male | 436 (56·4) | 123 (28·2) | 313(71·8) | 0·0007 |
| Female | 337 (43·6) | 134(39·8) | 203(60·2) |  |
| **Blood groups** |  |  |  |  |
| A+ | 142(20) | 49(34·5) | 93 (65·5) | 0·36 |
| A- | 3 (0·4) | 0 (0) | 3 (100) |  |
| AB+ | 86(12·1) | 26(30·2) | 60(69·8) |  |
| AB- | 4 (0·6) | 1 (25) | 3 (75) |  |
| B+ | 246(34·7) | 99(40·2) | 147(59·8) |  |
| B- | 16(2·3) | 5(31·3) | 11(68·8) |  |
| O+ | 199(28·1) | 61(30·7) | 138(69·3) |  |
| O- | 13 (1·8) | 3(23·1) | 10(76·9) |  |
| Don't know | 64 (8·3) | 13 | 51 |  |
| **BMI** |  |  |  |  |
| Normal or underweight ($\leq 24·9$) | 413 (53·4) | 154 (33·9) | 300 (66·1) | 0·21 |
| Overweight (25-29·9) | 248 (32·1) | 86 (34·7) | 162 (65·3) |  |
| Obese ($\geq 30$) | 71 (9·2) | 17 (23·9) | 54 (76·1) | |
| **Educational level** |  |  |  |  |
| Secondary and below | 60 (7·8) | 17(28·3) | 43 (71·7) | 0·13 |
| Higher secondary level | 121 (15·7) | 35(28·9) | 86(71·1) |  |
| Graduate | 354 (45·8) | 112(31·6) | 242(68·4) |  |
| Postgraduate | 233 (30·1) | 91(39·1) | 142(60·9) |  |
| Prefer not to specify | 5 (0·06) | - | - |  |
| **Workplace/occupation** |  |  |  |  |
| Hospital employee | 395 (51·1) | 156(39·5) | 239(60·5) | 0·0002 |
| Non-Hospital employee | 378 (48·9) | 101(26·7) | 277(73·3) |  |
| **Smoking/tobacco products consumption** |  |  |  |  |
| Yes | 134 (17·3) | 31 (23·1) | 103(76·9) | 0·006 |
| No | 639 (82·7) | 226(35·4) | 413(64·6) |  |
| **Alcohol consumption** |  |  |  |  |
| Yes | 229 (29·6) | 70(30·6) | 159(69·4) | 0·30 |
| No | 544 (70·4) | 187(34·4) | 357(65·6) |  |
| **Place of COVID-19 test conducted** |  |  |  |  |
| Government | 743 (96·1) | 249(33·5) | 494(66·5) | 0·43 |
| Private | 30 (3·9) | 8(26·7) | 22(73·3) |  |
| **Places of COVID-19 management** |  |  |  |  |
| Non-Hospital | 604 (78·2) | 206 (80·2) | 398(77·1) | 0·44 |
| Hospital | 169 (21·8) | 51(30·2) | 118(69·8) |  |
| **Severity of COVID-19 disease (Self-rated)** |  |  |  |  |
| Asymptomatic | 138 (17.8) | 21(15.2) | 117(84.8) | 0.0001 |
| Mild | 443 (57.3) | 154(34.8) | 289(65.2) |  |
| Moderate | 171 (22.1) | 71(41.3) | 101(58.7) |  |
| Severe | 21 (2.71) | 11(55) | 9(45) |  |
| **Management for COVID-19** |  |  |  |  |
| Managed with oxygen | 23 (2.98) | 15(65.2) | 8(34.8) | 0.0001 |
| Managed without Oxygen | 750 (97.0) | 242(32.3) | 508(67.7) |  |
| **Status of COVID-19 vaccination at the time survey** |  |  |  |  |
| Not vaccinated before COVID-19 infection | 407 (52.7) | 142(34.9) | 265(65.1) | 0.05 |
| 1<sup>st</sup> dose before COVID-19 virus infection | 175 (22.6) | 65(37.1) | 110(62.9) |  |
| 2<sup>nd</sup> dose before COVID -19 virus infection | 191 (24.7) | 50(26.5) | 141(73.5) |  |
| **Co-morbidities among participants** |  |  |  |  |
| Yes | 261 (33.8) | 110(42.1) | 151(57.9) | 0.0002 |
| No | 512 (66.2) | 147(28.7) | 365(71.3) |  |
| **Rating overall health status following COVID-19 illness** |  |  |  |  |
| Good Health | 709 (91.7) | 215(30.3) | 494(69.7) | 0.0001 |
| Poor or very poor health | 64 (8.3) | 42(65.6) | 22(34.4) |  |

The median age of the respondents was 34·0 (IQR 27·0-44·0) years. The proportion of males (56·4%) was slightly greater compared to female respondents. The body mass index was equal to or more than 25 for 41·3% (319/773) of the respondents. The most common reported blood group was B Rh positive (31·8%), followed by O Rh positive (25·7%). More than half of the respondents (51·1%) were currently employed in the hospital, and nearly 80% (587/773) of them were graduate and higher in education. A total of 17·3% (134/773) respondents smoked or chewed tobacco products and 29·6% (229/773) consumed alcohol (Table 1).

More than half of the respondents (52·7%, 407/773) were not vaccinated against the novel coronavirus and one quarter (24·7%) had received the second dose of the vaccine. About three fourths of participants (75·2%, 581/773) reported that they had either asymptomatic or mild acute COVID-19 disease and 2·71% (21) reported that they experienced severe acute COVID-19 illness (Table 1). Around 21·9% (169/773) of participants reported that they were managed in the hospital. Of these, 2·9% (23) required oxygen supplementation during the management. Further, 8·3% of participants considered themselves that they had poor or very poor overall health status following COVID-19 illness.

### Post-COVID-19 symptoms

One third of the participants (33·2%, 257/773) reported they had at least one or more ST-PCS (four weeks or more) but reduced to 12·8% (99/773) at 12 weeks or more and 0·90% (15/773) at 16 weeks or more since the SARS-CoV-2 tested positive. Few respondents (81) graded their severity of PCS as mild (58·0%, 47/81), moderate (34·6%, 28/81), and severe (4·93%, 4/81; Table 2). Whereas during acute phase of COVID-19 illness, 68·1% (175/257), 27·6% (71/257) and 4·2% (11/257) had an asymptomatic or mild, moderate and severe acute illness, respectively among participants who experienced PCS (Table 2). Another 80·2% (206/257) of the PCS respondents were not hospitalized during the acute COVID-19 illness (Table 1).

**Table 2:**
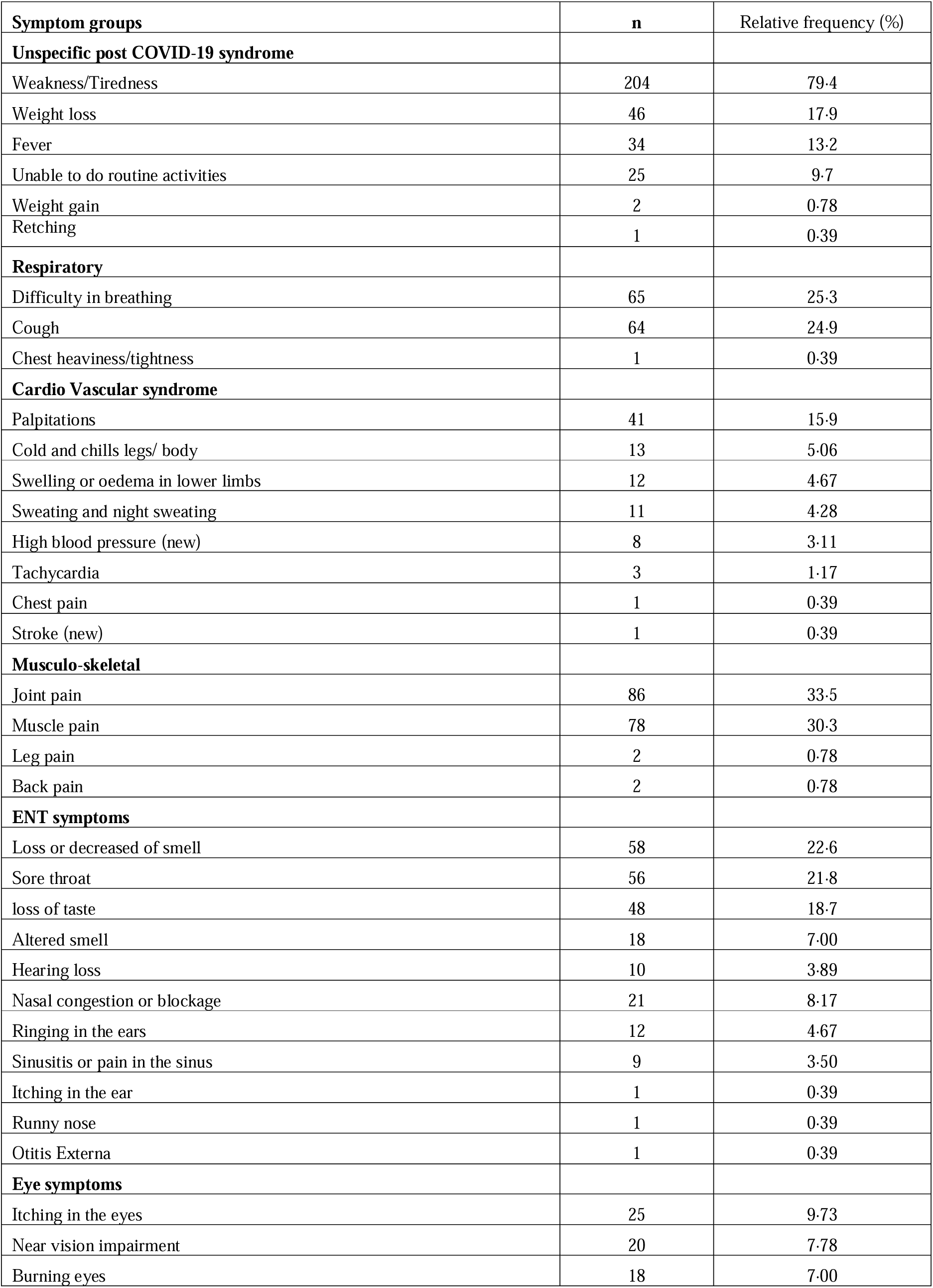
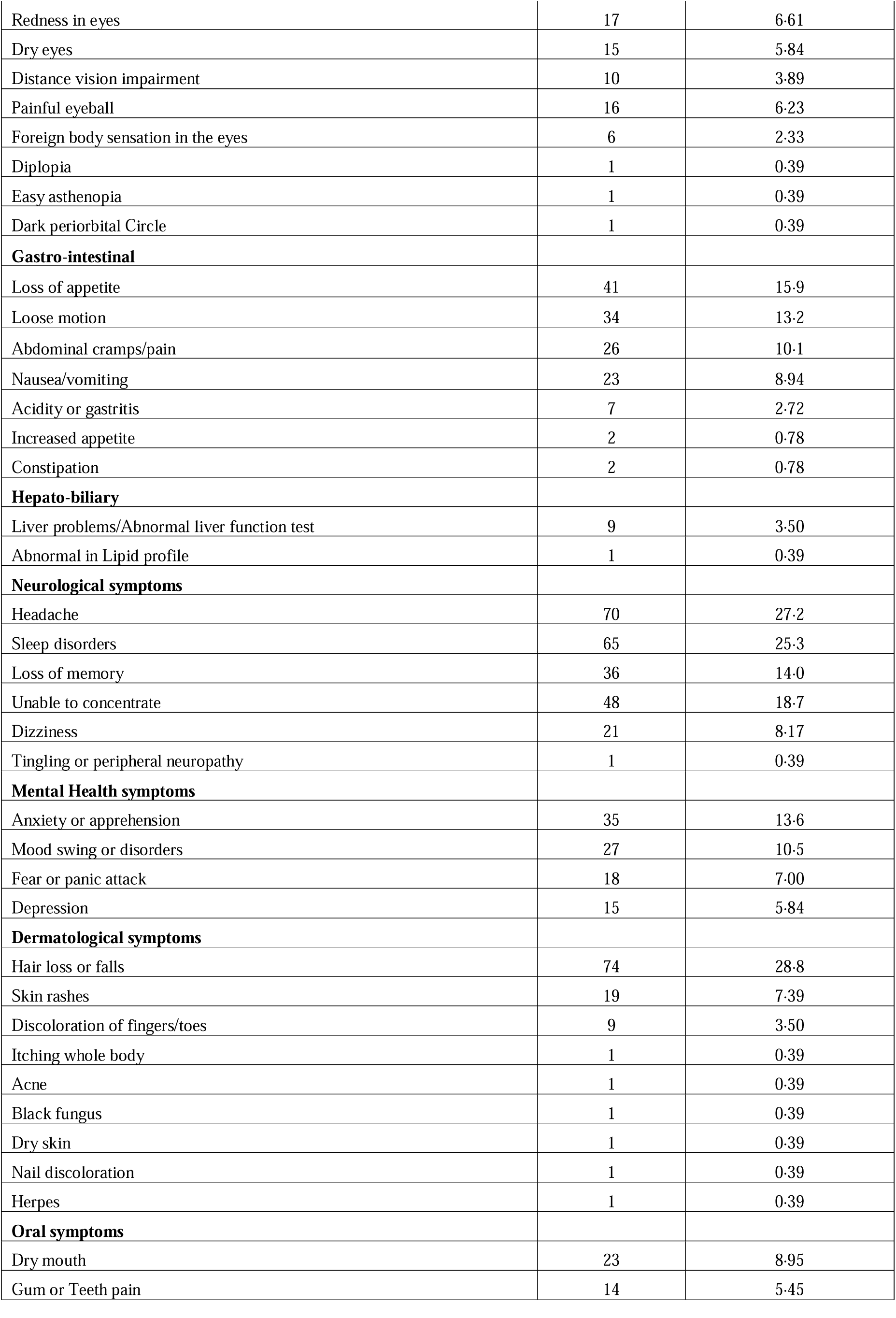
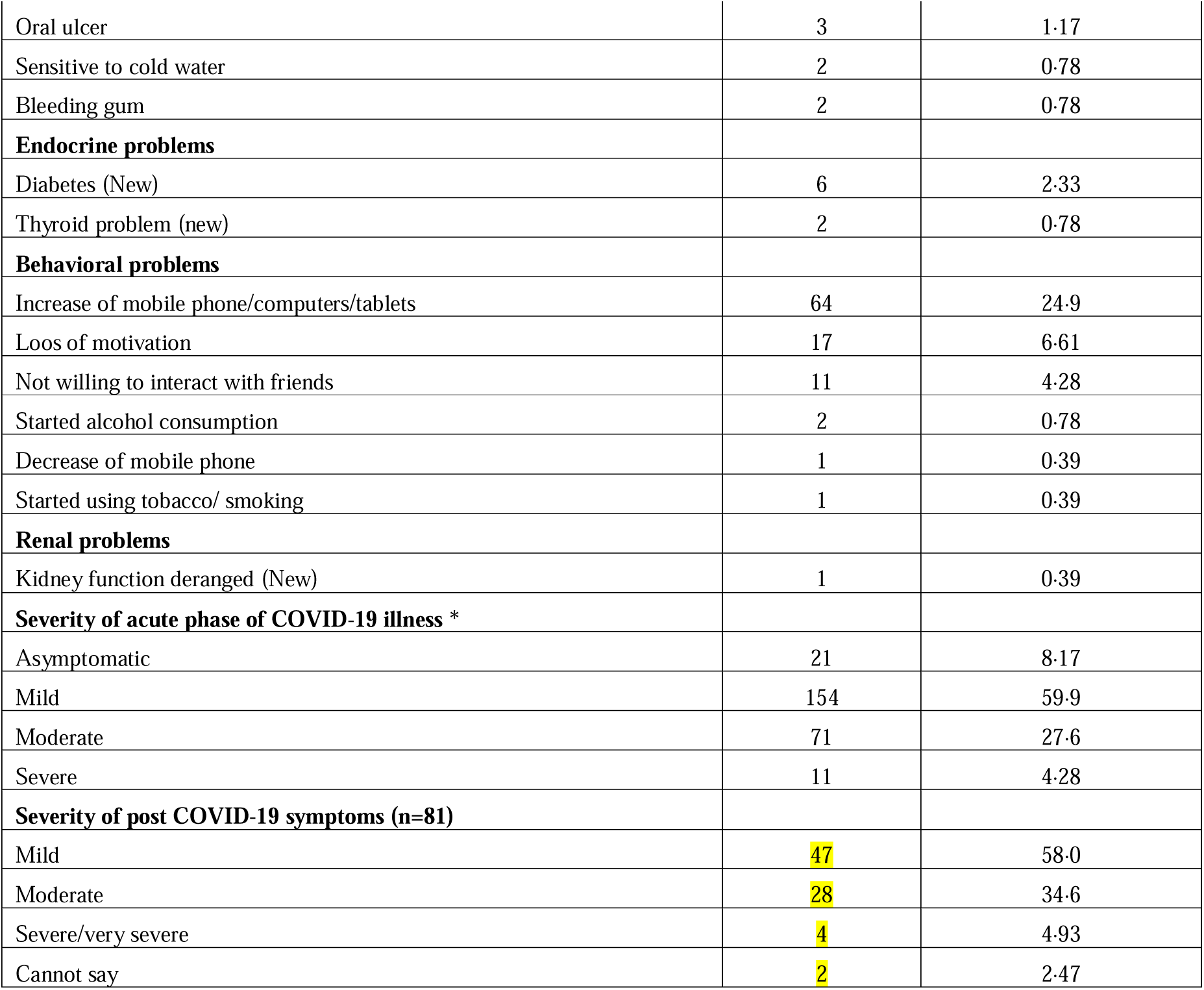
Characterization of post COVID-19 symptoms (n=257; 33·2% of total participants)

| Symptom groups | n | Relative frequency (%) |
| --- | --- | --- |
| **Unspecific post COVID-19 syndrome** |  |  |
| Weakness/Tiredness | 204 | 79.4 |
| Weight loss | 46 | 17.9 |
| Fever | 34 | 13.2 |
| Unable to do routine activities | 25 | 9.7 |
| Weight gain | 2 | 0.78 |
| Retching | 1 | 0.39 |
| **Respiratory** |  |  |
| Difficulty in breathing | 65 | 25.3 |
| Cough | 64 | 24.9 |
| Chest heaviness/tightness | 1 | 0.39 |
| **Cardio Vascular syndrome** |  |  |
| Palpitations | 41 | 15.9 |
| Cold and chills legs/ body | 13 | 5.06 |
| Swelling or oedema in lower limbs | 12 | 4.67 |
| Sweating and night sweating | 11 | 4.28 |
| High blood pressure (new) | 8 | 3.11 |
| Tachycardia | 3 | 1.17 |
| Chest pain | 1 | 0.39 |
| Stroke (new) | 1 | 0.39 |
| **Musculo-skeletal** |  |  |
| Joint pain | 86 | 33.5 |
| Muscle pain | 78 | 30.3 |
| Leg pain | 2 | 0.78 |
| Back pain | 2 | 0.78 |
| **ENT symptoms** |  |  |
| Loss or decreased of smell | 58 | 22.6 |
| Sore throat | 56 | 21.8 |
| loss of taste | 48 | 18.7 |
| Altered smell | 18 | 7.00 |
| Hearing loss | 10 | 3.89 |
| Nasal congestion or blockage | 21 | 8.17 |
| Ringing in the ears | 12 | 4.67 |
| Sinusitis or pain in the sinus | 9 | 3.50 |
| Itching in the ear | 1 | 0.39 |
| Runny nose | 1 | 0.39 |
| Otitis Externa | 1 | 0.39 |
| **Eye symptoms** |  |  |
| Itching in the eyes | 25 | 9.73 |
| Near vision impairment | 20 | 7.78 |
| Burning eyes | 18 | 7.00 |
| Redness in eyes | 17 | 6.61 |
| Dry eyes | 15 | 5.84 |
| Distance vision impairment | 10 | 3.89 |
| Painful eyeball | 16 | 6.23 |
| Foreign body sensation in the eyes | 6 | 2.33 |
| Diplopia | 1 | 0.39 |
| Easy asthenopia | 1 | 0.39 |
| Dark periorbital Circle | 1 | 0.39 |
| **Gastro-intestinal** |  |  |
| Loss of appetite | 41 | 15.9 |
| Loose motion | 34 | 13.2 |
| Abdominal cramps/pain | 26 | 10.1 |
| Nausea/vomiting | 23 | 8.94 |
| Acidity or gastritis | 7 | 2.72 |
| Increased appetite | 2 | 0.78 |
| Constipation | 2 | 0.78 |
| **Hepato-biliary** |  |  |
| Liver problems/Abnormal liver function test | 9 | 3.50 |
| Abnormal in Lipid profile | 1 | 0.39 |
| **Neurological symptoms** |  |  |
| Headache | 70 | 27.2 |
| Sleep disorders | 65 | 25.3 |
| Loss of memory | 36 | 14.0 |
| Unable to concentrate | 48 | 18.7 |
| Dizziness | 21 | 8.17 |
| Tingling or peripheral neuropathy | 1 | 0.39 |
| **Mental Health symptoms** |  |  |
| Anxiety or apprehension | 35 | 13.6 |
| Mood swing or disorders | 27 | 10.5 |
| Fear or panic attack | 18 | 7.00 |
| Depression | 15 | 5.84 |
| **Dermatological symptoms** |  |  |
| Hair loss or falls | 74 | 28.8 |
| Skin rashes | 19 | 7.39 |
| Discoloration of fingers/toes | 9 | 3.50 |
| Itching whole body | 1 | 0.39 |
| Acne | 1 | 0.39 |
| Black fungus | 1 | 0.39 |
| Dry skin | 1 | 0.39 |
| Nail discoloration | 1 | 0.39 |
| Herpes | 1 | 0.39 |
| **Oral symptoms** |  |  |
| Dry mouth | 23 | 8.95 |
| Gum or Teeth pain | 14 | 5.45 |
| Oral ulcer | 3 | 1.17 |
| Sensitive to cold water | 2 | 0.78 |
| Bleeding gum | 2 | 0.78 |
| **Endocrine problems** |  |  |
| Diabetes (New) | 6 | 2.33 |
| Thyroid problem (new) | 2 | 0.78 |
| **Behavioral problems** |  |  |
| Increase of mobile phone/computers/tablets | 64 | 24.9 |
| Loss of motivation | 17 | 6.61 |
| Not willing to interact with friends | 11 | 4.28 |
| Started alcohol consumption | 2 | 0.78 |
| Decrease of mobile phone | 1 | 0.39 |
| Started using tobacco/ smoking | 1 | 0.39 |
| **Renal problems** |  |  |
| Kidney function deranged (New) | 1 | 0.39 |
| **Severity of acute phase of COVID-19 illness \*** |  |  |
| Asymptomatic | 21 | 8.17 |
| Mild | 154 | 59.9 |
| Moderate | 71 | 27.6 |
| Severe | 11 | 4.28 |
| **Severity of post COVID-19 symptoms (n=81)** |  |  |
| Mild | 47 | 58.0 |
| Moderate | 28 | 34.6 |
| Severe/very severe | 4 | 4.93 |
| Cannot say | 2 | 2.47 |

Among individuals having post COVID-19 symptoms, a large proportion (85·6%, 220/257) reported unspecific post covid-19 symptoms (UPCS, table 2). This was followed by musculoskeletal manifestations (49·8%, 128/257), Ear Nose and Throat symptoms (47·5%), neurological (47·0%, 121/257), cardio-respiratory (42·4%, 109/257), gastrointestinal (36·2%, 93/257), ocular symptoms (31·9%), dermatological symptoms (31·5%, 81/257), and cardio-vascular (24·5%, 63/257) symptoms, and mental health symptoms (23·7%). The remaining organ system specific symptoms, including behavioral changes, oral health problems, hepatobiliary, endocrine, kidney function abnormality were reported by less than 20% of the respondents (Figure 1). All in all, the most commonly reported symptoms were fatigue, pain in the joins and muscle, hair loss and headache, cough, breathlessness, sleep disorders, sore throat and decrease of smell and taste (Figure 2).

**Figure 1:**
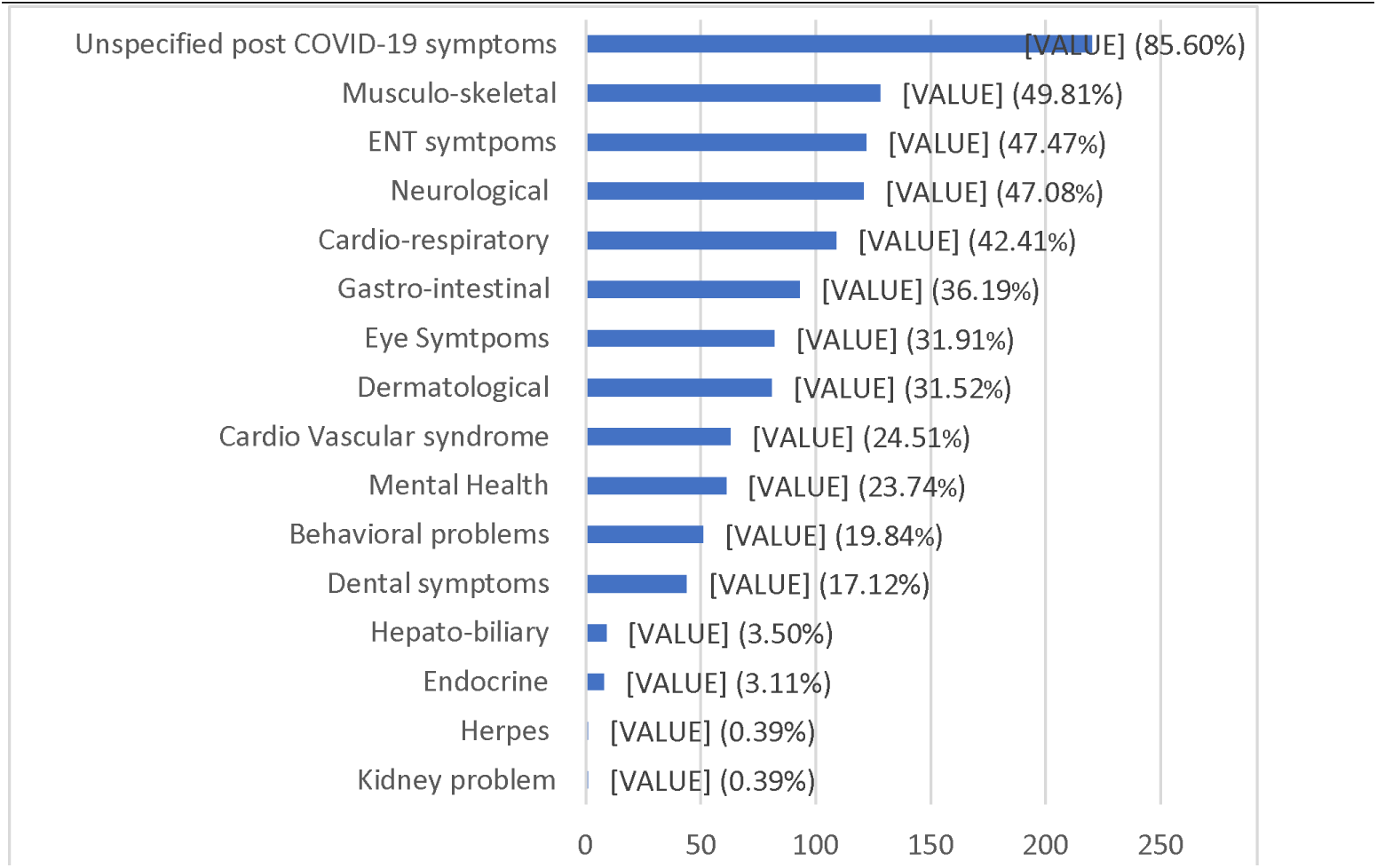
Organ system specific post COVID-19 manifestations (Counts and relative frequency in bracket)

**Figure 2:**
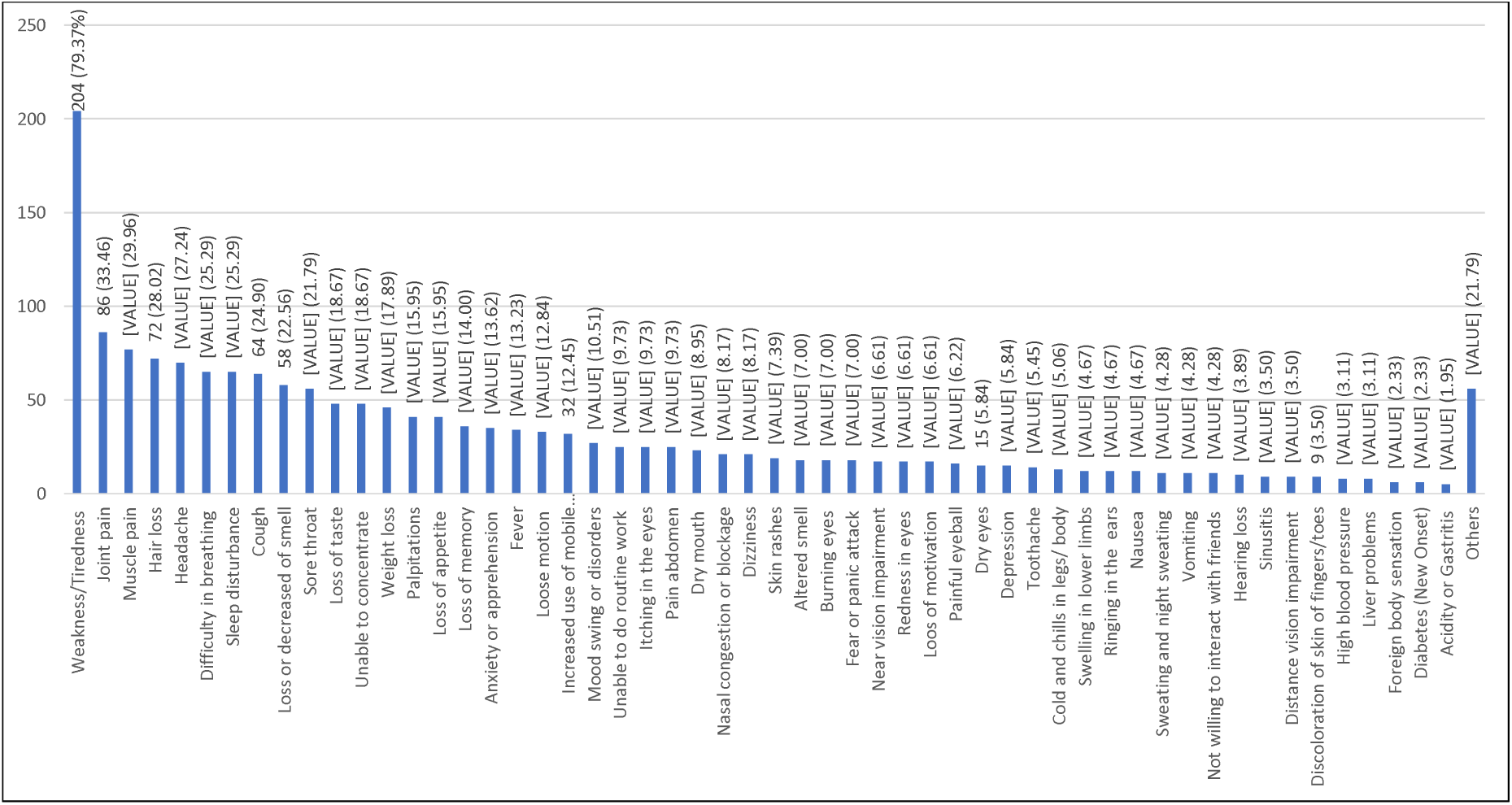
Post COVID-19 manifestations in counts and relative frequency (round bracket)

### Pre-existing co-morbidities among participants

Among the participants, 33·8% (261/773) reported that they had at least one or more associated co-morbidities before the COVID-19 disease (Figure 3). Hypertension (10·1%, 78/773) was the most common co-morbid, followed by diabetes mellitus (6·33%, 49/773). Other reported common co-morbid conditions were thyroid problems (3·8%, 29/773), migraine (3·6%, 28/773), heart disease and asthma each as 2·46% (19/773), etc. The remaining co-morbidities were present in less than 2% of participants. (Figure 3).

**Figure 3:**
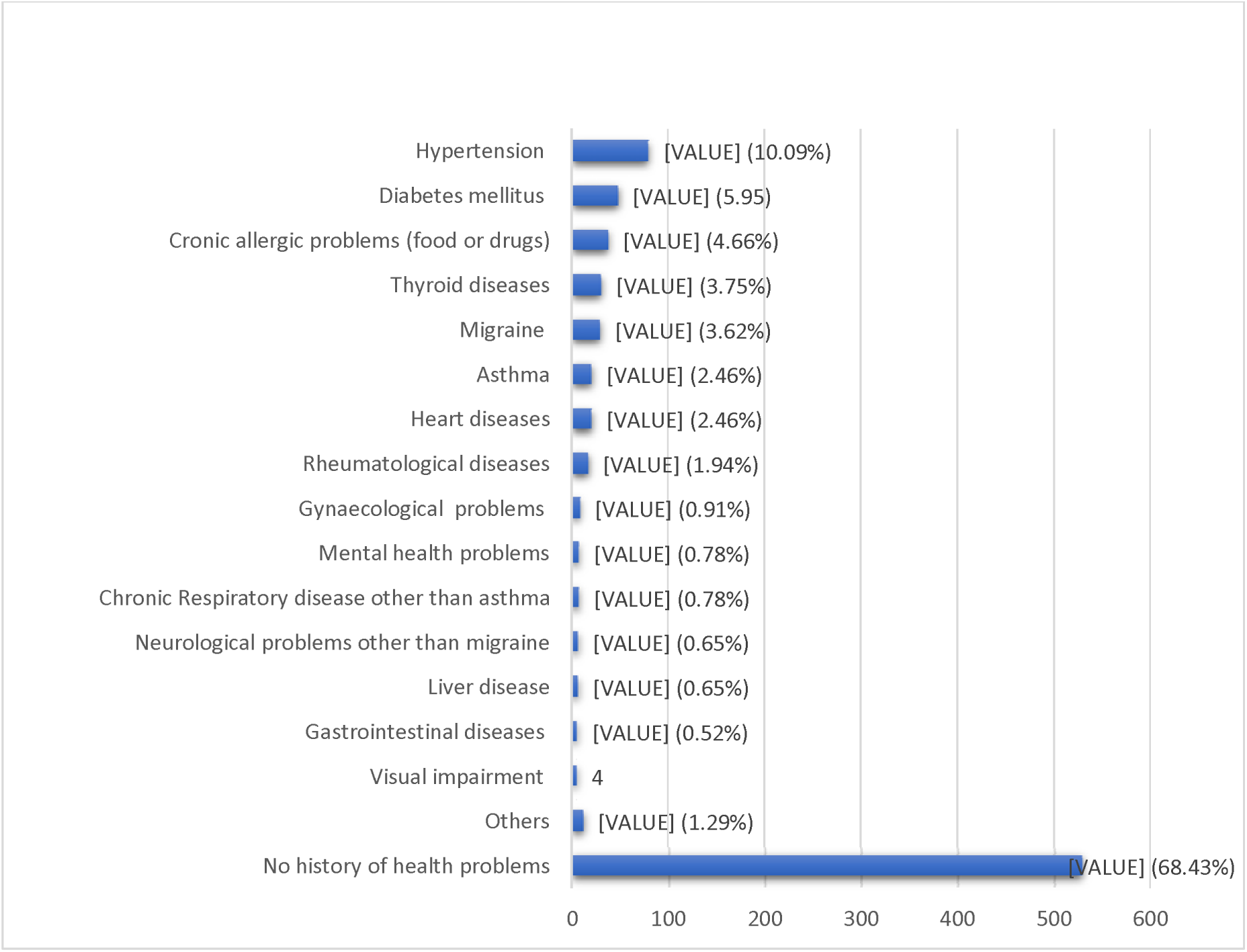
Co-morbidities associated among study participants (counts and relative frequency)

### Factors associated with Post COVID-19 manifestations

The analysis (Table 1) showed that the Post COVID-19 symptoms were significantly varied across the categorical age groups (p< 0·0001), gender (p<0·0007), place of work (p<0·0002), severity of COVID (p<0·0001), oxygen supplementation during the treatment (p<0·0001), individuals with vaccination status (p<0·05), pre-existing co-morbidities (p<0·0002). No association was found with blood groups and post COVID-19 symptoms and body mass index.

The univariable logistic regression model showed that several factors were associated with post COVID-19 manifestations (Table 3). Younger individuals (p-value <0·001; Odds ratio (OR) 2·93; 2·10; 6·62; 4·73 for categorical age groups 26-45; 26-60; 61-70; more than 70 years of age), male gender (OR 0·59; 95% CI 0·4-0·8), individuals working other than hospital (OR 0·55; 95% CI 0·41-0·75) and received second dose of COVID-19 vaccine (OR 0·65; 95% CI 0·45-0·96) have a lower risk for post COVID-19 symptoms. Oxygen supplementation during the treatment (OR 3·93; 95%CI 1·64-9·40), severity of COVID-19 illness (OR 2·96; 95%CI 1·79-4·91; OR 3·91; 95%CI 2·24-6·82; OR 6·80; 95%CI 2·51-18·4 for mild, moderate and severe cases) are all associated with PCS.

**Table 3:**
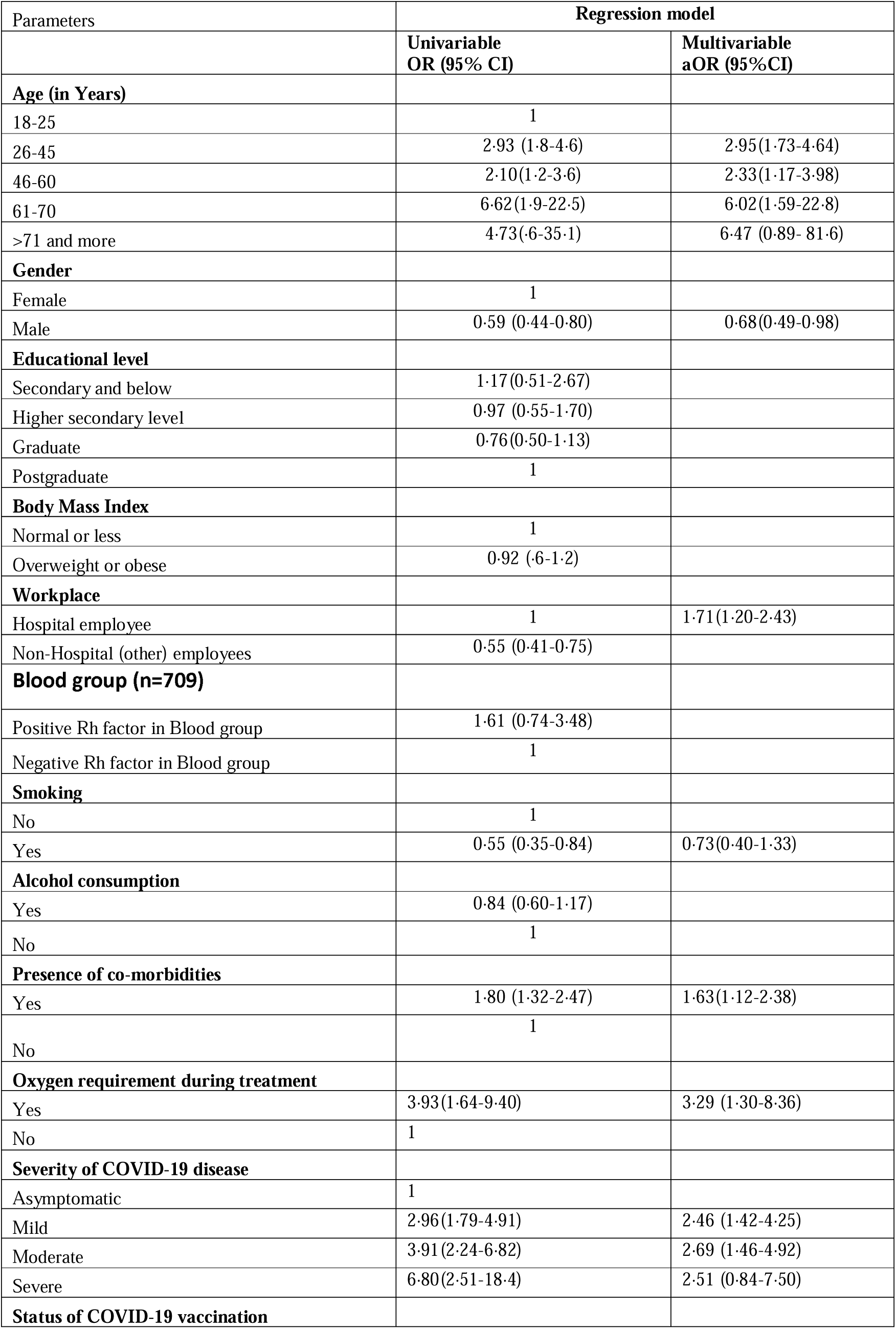
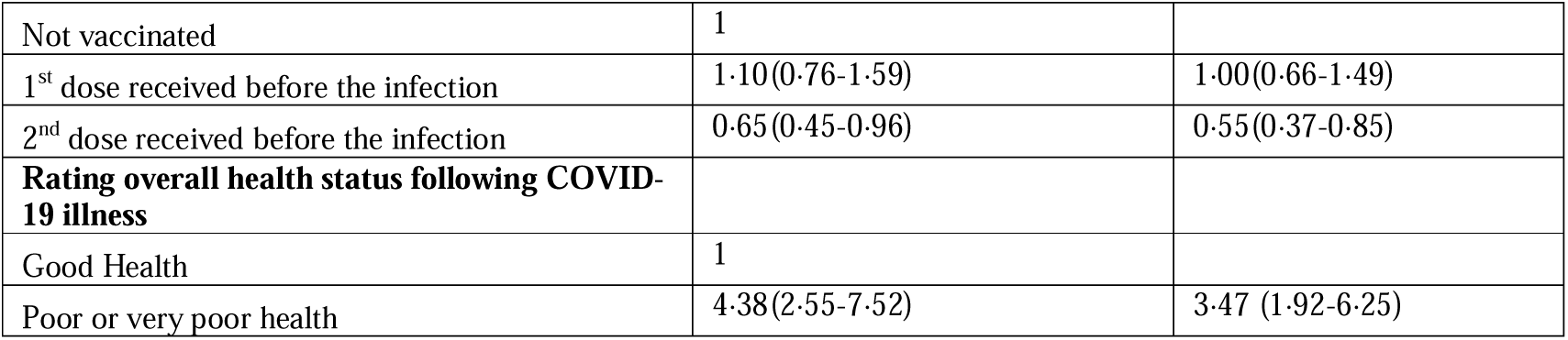
Factors associated with post COVID-19 symptoms.

The multivariate analysis shows that PCS are more likely to be developed at 1·63 times in pre-existing co-morbid than healthy participants; 3·29 times in patients with oxygen supplementation during the treatment; 2·46 to 6·8 times in mild to severe COVID-19 illness than asymptomatic patients. Smoker compared to non-smokers was found to be a protective in univariable analysis, but in the multivariable analysis, after adjusting other explanatory variables, smoking was not associated with post COVID-19 symptoms.

One of the important findings in the multivariable regression model was odds of developing post COVID-19 symptoms was lower among individuals who have received second dose of COVID-19 vaccine when compared with unvaccinated individuals (OR 0·55; 95%CI 0·37-0·85, Table 3). All in all, the multivariable logistic regression model revealed that older age groups, female gender, hospital employees, oxygen supplementation at the COVID-19 management, cognitive or memory impairment, severity of acute COVID-19 illness, persons who have not vaccinated were independent risk factors for prolonged COVID-19 manifestations (Table 3)

## Discussion

In our study, the participants included were tested positive for SARS-CoV-2 at least four weeks or more before the date of inception of data collection. We also enquired about the status of COVID-19 illness during the telephonic calls to ensure the information we collected was for post COVID-19 manifestations. As of now, it is practically challenging to compare the findings and the prevalence of PCS manifestations across various studies due to differences in assessment time since recovery, and variability in the duration while defining post COVID-19 symptoms. Therefore, the prevalence of long COVID, to date, ranges from 27·8% to 95·0% depending on the collection of data time since recovery. ^6,18,19,20^

The present study shows that the prevalence of PCS was 33·2%, both ST and LT-PCS. This indicates that one in three individuals have persistent PCS after four weeks or more following positive test. Further, it is evident that many patients continue to experience persistent symptoms after COVID-19, irrespective of disease severity during acute illness and hospitalization. Studies have reported multiple PCS in non-hospitalized patients after several months of recovery from COVID-19.^21,22^ Though individuals reporting the severity of PCS were less in number, the study shows PCS range from mild to severe.

In our study, the prevalence of PCS was slightly higher in males (56·4%) compared to females (43·6%), but female gender is more likely to develop PCS than males. The difference in physiological and social factors or physical activities may be a possible explanation for this, but further study is warranted to rule out. Female predominant for PCS is also reported in other studies. ^21,23^ The present study also indicates that older age individuals are more likely to develop PCS compared to younger age groups (Table 1 and 3). Several studies also noted that increasing age is a risk factor for PCS. ^18,24,19^ Education, alcohol consumption, body mass index, and blood groups were not revealed to have any significant effect on the development of PCS.

The study noted that individuals working in the healthcare sectors have a higher risk of developing PCS compared to individuals who are working in other sectors. In the multivariable regression analysis, that the hospital employee is 1·7 times higher risk of having PCS than their counterpart non-hospital employees (Table 3). Participants with more severe COVID-19 illness are likely to be experienced a higher risk for PCS after recovery. However, a longitudinal follow up study would be required to assess the duration of persistence for PCS among severe acute illness participants in the future.

In the present study, fatigue was the most common among all PCS, reaching up to 80·3%. Other studies conducted elsewhere, including systemic review and meta analysis, fatigue was the most frequently reported symptom as of today with the prevalence ranging from 30% to 82·9%. ^20,25,21 26^ The remaining, such as pain in joints and muscle, hair loss, headache, shortness of breath, sleep disturbance, cough, loss of smell, and taste were noted in approximately 20 to 34% of participants. These findings are consistent with other studies as among frequently reported PCS. The pathophysiology behind such a wide range manifestation is not yet cleared, however, it indicates the multiorgan involvement as in the acute phase of the illness. The possible immunological mechanism involving the multi-organ system following SARS-CoV-2 infection is explained for the appearance of these long lasting symptoms in other literature. ^27,20^

The present study also shows that the odds of having PCS is reduced by 45 % in individuals who have received double dose of COVID-19 vaccine compared to persons who have not received any dose. This is an additional beneficial finding for COVID-19 vaccine that has already reduced the risk of SARS CoV-2 infection and the severity of acute illness. Therefore, vaccination against COVID-19 should be encouraged among the eligible population as early as possible.

Few percentages of recovered patients also experienced both near and distance visual impairment, and dry eyes. It is not sure whether symptoms could be side effects of medication that was commonly used during the acute management, e.g., hydroxychloroquine. Since the post covid assessment was done after four or more weeks of recovery, symptoms like red and painful eyes could be due to immunological mechanisms, but it needs further study. The study also indicates a strong association between the pre-existing co-morbid and the presence of PCS. Similar pre-existing co-morbid such as hypertension, chronic respiratory diseases, diabetes mellitus are shown to be determinants of the prolonged COVID-19 symptoms as illustrated in other studies.^24 28,29^ Further, our study shows that participants who had managed the acute phase of COVID-19 with oxygen supplementation are more likely to develop PCS compared to participants managed without oxygen. A similar finding is also indicated in a study conducted elsewhere.^30^

Among psychosocial behavioral changes, a sizeable number of recovered subjects also reported anxiety, mood disorders, panic and depression in our study. Various changes in behavioral aspects in the present study were noted, such as a substantial increase in the use of electronic gadgets among the survivors. Such a link with mental and behavioral problems among recovered patients is also reported in other studies. ^28,5,31^

All in all, we observed that the present study clearly indicates that PCS involves almost all organ systems of the body. Such a multi-organ involvement in PCS has been described in the literature. ^5,32^ Similar findings were reported in a mobile phone app-based study in the United Kingdom.^33^ This implies managing such wide varied symptoms needs a holistic multi-disciplinary approach involving multi-specialties, including hospital and community-based rehabilitation program to support the healthcare and psychological needs for an extended time rather than organ specific management.

To date, it is not yet fully understood how long the PCS will continue to persist among COVID-19 survivors. In addition to our findings, various previous studies reported that SARS-CoV-2 can still infect individuals despite vaccination, though the severity of the disease can be reduced. Moreover, we are not sure that how long the ongoing wave of the COVID-19 will continue to stay with us. PCS can also affect individuals regardless of the severity of the acute phase of COVID-19 illness. Therefore, there is an urgent need of developing a PCS care model which is suitable for the resource limited countries. One such model is being run to manage PCS in the United Kingdom. ^34^ Individuals with PCS who have pre-existing co-morbid would be required a proper follow up strategy because such persons are the potential candidates for developing significant disability and impairment in their daily functioning. In addition, a few patients who had recovered from COVID-19 also had newly detected diabetes, abnormal renal, liver, and thyroid function tests and stroke. Similar findings are also shown in other studies.^24^ These patients need to be investigated thoroughly per se, including those who reported poor or very poor health following COVID-19 and managed accordingly. At the same time, follow up is necessary to know whether such conditions could be reversible in due course of time. Finally, a significant number of participants were reported of having poor or very poor health following COVID-19 diseases.

The limitations of our study include that our study may not be truly representative of all communities since we covered those who are EHS beneficiaries of one institute, and we also could not collect data from the participants who were not using either smartphones or WhatsApp, despite we requested alternative numbers that had the WhatsApp. We were not able to assess the reverse casualty in some of PCS and their association. For examples, pain in joints is due to PCS or pre-existing joint problems.

## Conclusion

The PCS affect a wide range of body organ systems, regardless of the severity of the acute phase of illness. Female gender, older individuals, oxygen supplementation during the acute illness, associated co-morbidities, severe acute illness are the key predictors of PCS. Such persistent COVID-19 manifestations are not only a burden to the affected individuals and their families but also challenges of healthcare and public health service. We suggest an integrated care model involving all relevant healthcare disciplines while managing PCS in the outpatient setting at every healthcare facility, rather than organ-specific approaches. Prioritization of the follow-up care may be given for those with pre-existing co-morbid and elder people. Interestingly, our finding highlights the importance of vaccination in the reduction of PCS and shows that the odds of having prolonged COVID-19 symptoms are cut in individuals who have received two doses of the COVID-19 vaccine compared to unvaccinated individuals.

It is not yet fully understood that how long these symptoms will persist in recovery individuals. Continuous follow-up will be important to assess the further prolonged post COVID-19 health problems. At the same time, a community-based rehabilitation program, including psychological support should be a part of post COVID-19 care. The strength of our study lies first, our team telephonically contacted all participants that ensure not only enquire on COVID-19 illness status or recovery but also to avoid the potential of multiple responses. Second, the study population is either employees of our hospital or their dependents who help the accuracy in reporting PCS. Third, the team can contact for follow up in line with a longitudinal study in the future.

## Data Availability

The post COVID-19 data collected for this study, including anonymized participants information will be made available publicly in the interest of public health research. Interested researchers and participants can contact the corresponding author (SSS).

## Acknowledgment

We would like to thank all respondents who are employees of All India Institute of Medical Sciences New Delhi and their respective dependents for their participation in the post COVID-19 survey. We thank all the staff who called telephonically to the participants since from day one of the survey. We appreciate Mr. Vikas for his relentless support in converting the questionnaires in the electronic format using Google forms and assistance in data cleaning. We also like to thank all members in ethics committee for their quick approval of the study and staff who handled the institute database. Finally, we appreciate the staff working in database management of All India Institute of Medical Sciences New Delhi for their support in providing the list of participants for the study.

## Author contributions

SSS conceived the study and designed the survey with support from KA, TJS, TR, and GR. SS, GR, VP were responsible for data collection. SR, SSS, MS, GV accessed and cleaned the preliminary raw data. SR, MS, SSS planned and performed the statistical analysis. SSS created tables with inputs from YPSB, MK. SSS, MS created the figures. SSS, YPSB, MS, NN, YG, AR, GN designed the study instrument. SSS wrote the manuscript, and edited by the draft from YPSB. PK, AHN, PV, DL helped in procuring the list of COVID-19 survivors of the institute. All authors listed meet authorship criteria, and reviewed the manuscript before finalization and had full access the data. SSS is the principal investigator and guarantor.

## References

1. World Health Organization G· WHO Director-General’s opening remarks at the media briefing on COVID-19 - 11 March 2020. (Accessed on August 21, 2021)

2. Zhu N, Zhang D, Wang W, et. al. A novel coronavirus from patients with pneumonia in China, 2019. N Engl J Med 2020;382:727–33.

3. World Health Organization G. WHO Coronavirus (COVID-19) Dashboard, WHO Coronavirus (COVID-19) Dashboard With Vaccination Data. (Accessed on August 21, 2021).

4. The Government of India. IndiaFightsCorona COVID-19 in India, Corona Virus Tracker mygov.in. https://www.mygov.in/covid-19 (accessed on August 19, 2021).

5. Nalbandian A, Sehgal K, Gupta A, et al. Post-acute COVID-19 syndrome. Nat Med 2021;27:601–15.

6. Carfí A, Bernabei R, Landi F. Persistent symptoms in patients after acute COVID-19. JAMA - J Am Med Assoc 2020;324:603–5.

7. Garrigues E, Janvier P, Kherabi Y, et al. Post-discharge persistent symptoms and health-related quality of life after hospitalization for COVID-19. J Infect 2020;81:e4–6.

8. Mandal S, Barnett J, Brill SE, et al. Long-COVID’: A cross-sectional study of persisting symptoms, biomarker and imaging abnormalities following hospitalisation for COVID-19. Thorax 2021;76:396–8.

9. Nehme M, Braillard O, Alcoba G, et al. COVID-19 Symptoms: Longitudinal Evolution and Persistence in Outpatient Settings. Ann Intern Med 2021;174:723–5.

10. Greenhalgh T, Knight M, A’Court C, Buxton M, Husain L. Management of post-acute covid-19 in primary care. BMJ 2020;370.

11. Google Forms: Free Online Surveys for Personal Use. https://www.google.com/forms/about/

12. Karagöz MA, Gül A, Borg C, et al. Influence of COVID-19 pandemic on sexuality: a cross-sectional study among couples in Turkey. Int J Impot Res December 2020:1–9.

13. Gondwal R, Pal A, Paul S, Bohra R, Pal A, Aulakh S, Bhat A. Sexual Behavior During the Times of COVID-19-Related Lockdown in India: Results From an Online Survey. J Psychosexual Heal 2020;2:242–6.

14. Singer B, Walsh CM, Gondwe L, Reynolds K, Lawrence E, Kasiya A. WhatsApp as a medium to collect qualitative data among adolescents: lessons learned and considerations for future use. Gates Open Res 2020 4130 2020;4:130.

15. Arroz JAH, Candrinho BN, Mussambala F, Chande M, Mendis C, Dias S, Martins MDRO. WhatsApp: A supplementary tool for improving bed nets universal coverage campaign in Mozambique. BMC Health Serv Res 2019;19.

16. Kumar N, Sharma S. Survey Analysis on the usage and Impact of Whatsapp Messenger. Glob J Enterp Inf Syst 2017;8:52.

17. Eysenbach G. Improving the quality of web surveys: The Checklist for Reporting Results of Internet E-Surveys (CHERRIES). J Med Internet Res 2004;6.

18. Raveendran AV, Jayadevan R, Sashidharan S. Long COVID: An overview. Diabetes Metab Syndr 2021;15:869–75.

19. Iqbal A, Iqbal K, Arshad Ali S, et al. The COVID-19 Sequelae: A Cross-Sectional Evaluation of Post-recovery Symptoms and the Need for Rehabilitation of COVID-19 Survivors. Cureus 2021;13.

20. Nalbandian A, Sehgal K, Gupta A, et al. Post-acute COVID-19 syndrome. Nat Med 2021 274 2021;27:601–15.

21. Augustin M, Schommers P, Stecher M, et al. Post-COVID syndrome in non-hospitalised patients with COVID-19: a longitudinal prospective cohort study. Lancet Reg Heal Eur 2021;6:100122.

22. Heightman M, Prashar J, Hillman TE, et al. Post-COVID assessment in a specialist clinical service: a 12-month, single-centre analysis of symptoms and healthcare needs in 1325 individuals. medRxiv June 2021:2021.05.25.21257730.

23. Hossain MA, Hossain KMA, Saunders K, et al. Prevalence of Long COVID symptoms in Bangladesh: A Prospective Inception Cohort Study of COVID-19 survivors. medRxiv July 2021:2021.07.03.21259626.

24. Kamal M, Abo Omirah M, Hussein A, Saeed H. Assessment and characterisation of post-COVID-19 manifestations. Int J Clin Pract 2021;75.(3):e13746. doi: 10.1111/ijcp.13746. Epub 2020 Nov 3. PMID: 32991035; PMCID: PMC7536922.

25. Moreno-Pérez O, Merino E, Leon-Ramirez JM, et al. Post-acute COVID-19 syndrome. Incidence and risk factors: A Mediterranean cohort study. J Infect 2021;82:378–83.

26. Fahad M I, Kyle L, Viknesh S, Jonathan MC, Hutan A, Ara D. Characteristics and predictors of acute and chronic post-COVID syndrome: A systematic review and meta-analysis. EClinicalMedicine 2021;36.6, 2021.

27. Ramakrishnan RK., Kashour T, Hamid Q, Halwani R, Tleyjeh IM. Unraveling the Mystery Surrounding Post-Acute Sequelae of COVID-19. Front Immunol 2021 Jun 30;12:686029. doi: 10.3389/fimmu.2021.686029. PMID: 34276671; PMCID: PMC8278217.

28. Galal I, Hussein A, Amin MT, et al. S Determinants of persistent post-COVID-19 symptoms: value of a novel COVID-19 symptom score. Egypt J Bronchol 2021;15:1–8.

29. Fatima G, Bhatt D, Idrees J, Khalid B, Mahdi F, Mehdi F. Elucidating Post-COVID-19 manifestations in India. https://www.medrxiv.org/content/10.1101/2021.07.06.21260115v1 doi: https://doi.org/10.1101/2021.07.06.21260115

30. Rao GV, Gella V, Radhakrishna M, et al. Post-COVID-19 symptoms are not uncommon among recovered patients-A cross-sectional online survey among the Indian population. https://www.medrxiv.org/content/10.1101/2021.07.15.21260234v1

31. Amin-Chowdhury Z, Harris RJ, Aiano F, et al. Characterising post-COVID syndrome more than 6 months after acute infection in adults; prospective longitudinal cohort study, England. medRxiv April 2021:2021.03.18.21253633.

32. Yan, Z.; Yang, M.; Lai, C.-L. Long COVID-19 Syndrome: A Comprehensive Review of Its Effect on Various Organ Systems and Recommendation on Rehabilitation Plans. Biomedicines 2021, 9, 966. https://doi.org/10.3390/biomedicines9080966

33. Antonelli M, Penfold RS, Merino J, et al. Risk factors and disease profile of post-vaccination SARS-CoV-2 infection in UK users of the COVID Symptom Study app: a prospective, community-based, nested, case-control study. Lancet Infect Dis 2021;0.

34. Parkin A, Davison J, Tarrant R, et al. A Multidisciplinary NHS COVID-19 Service to Manage Post-COVID-19 Syndrome in the Community. J Prim Care Community Health 2021 Jan-Dec;12:21501327211010994. doi: 10.1177/21501327211010994. PMID: 33880955; PMCID: PMC8064663.PMC8064663.

